# DEVELOPMENT AND VALIDATION OF THE SRI LANKAN PRIMARY CARE ASSESSMENT TOOL (SL-PCAT) FOR EVALUATING PATIENTS’ EXPERIENCES IN PRIMARY CARE

**DOI:** 10.64898/2026.02.16.26346399

**Authors:** Parami Abeyrathna, Suneth Buddhika Agampodi, Shyamalee Samaranayake, Pahala Hangidi Gedara Janaka Pushpakumara

## Abstract

**Background:** This study developed and validated a primary care assessment tool (SL-PCAT) for evaluating patients’ experiences in the Sri Lankan healthcare setting.

**Methods:** The study was conducted from April 2021 to August 2023 inside a medical faculty and selected communities in the Anuradhapura District. The tool was validated through item development, scale development, and scale evaluation processes. Items were selected through focused-group interviews, a literature review, and an expert review. Two language experts translated the tool into the Sinhala language. The scale’s construct was evaluated parallel to a cross-sectional household survey in 32 villages. First, 633 valid data were randomly split into two groups (Subsample-1 [n=320] and Subsample-2 [n=313]). Exploratory factor analysis was performed in Subsample-1 with principal component analysis (PCA), followed by reliability assessment with Cronbach’s alpha. A confirmatory factor analysis (CFA) was performed on Subsample-2. Likert scale assumption testing and descriptive statistics were performed in the final factor structure.

**Results:** The four factor solution from PCA explained 66.8% variance in data. Domains identified were ‘contextual care at family and community with continuity’, ‘accessibility’, ‘patient-centred care’, and ‘comprehensive and coordinated care’. The domains reported Cronbach’s alpha values between 0.719 and 0.859. The goodness of fit index of the CFA improved after allowing a few modifications (X2=199, p<0.001, RMSEA= 0.079, GFI=0.916, SRMR=0.066, CFI=0.916). The corresponding convergent and discriminant validity of the Likert scale assumptions was optimum, with Item-scale correlations between 0.346 and 0.830 and scaling success rates over 70%. Three domains, except ‘contextual care at family and community with continuity’, reported positively skewed scores, indicating more positive responses.

**Conclusions:** SL-PCAT is a feasible, valid, and reliable tool for assessing patients’ experiences of primary care in Sri Lanka.

## INTRODUCTION

Primary care is a key healthcare process that provides first contact, accessible, coordinated, and continuous care addressing a wider range of patient’s healthcare needs (1). Primary care has evolved prominently within Sri Lanka’s solid primary healthcare system following its worldwide recognition at the Alma-Ata declaration in 1978 (2). Regular primary care assessments improve health outcomes by monitoring key healthcare processes such as managing leading non-communicable diseases (NCDs) and screening (3,4). Assessing the healthcare needs and outcomes of the patients is essential for efficient resource allocation and developing policies targeting interventions and health programs that address the population’s specific needs (5). Improving the quality of care by monitoring the performance of healthcare providers needs to be processed through provider- and patient-centred assessments (3,6).

Commonly utilised primary care assessment methods evaluate various service points from doctor-patients to higher-ups in the health system (7). These tools have been cross-validated for various languages, countries, and age groups, adjusting to unique features of individual health systems (8–10). Except for the primary care assessment tools in Tibet, South Korea and a few other versions, there are a few patients-based assessments in the Asian region, and none were found in the South Asian region. Because of socio-demographic factors, the organisation of the health system and cultural backgrounds, it is challenging to develop universal tools in the region.

Sri Lanka, a South Asian country, stands out for its better maternal and child health indicators and universal health coverage indices (11,12). The primary healthcare system, a dual model, functions through state and private sector stakeholders. New healthcare reforms have been introduced by the Ministry of Health, Sri Lanka, to align the prevailing primary care system to achieve the population and health system demands (13). Approximately 50-60% of primary care is delivered from the state sector (14). In a non-hierarchical professional model, primary care is delivered through specified professionals, e.g. doctors of outpatient departments (OPD) in state and private hospitals, primary medical care units (PMCU), divisional hospitals, general practitioners (GPs) or private allopathic doctors and, family physicians (15). Most GPs work part-time in the private sector during alternative working hours while permanently employed at state hospitals. Both the state and private primary care facilities are governed by the Ministry of Health Sri Lanka through sub-departments and councils at varied administrative levels. Most quality improvement interventions in Sri Lanka are based on facility and provider surveys (3,6,13,16). However, there is no direct mechanism for assessing care based on patient experiences. This study aimed to develop and validate a tool for assessing patients’ perceptions of the quality of primary care delivered by primary care doctors in Sri Lanka: SL PCAT (Sri Lankan Primary Care Assessment Tool).

## METHODS

### Study design

The study was conducted from April 2021 to August 2023 in the Anuradhapura District of Sri Lanka. The tool development was conducted as a cross-sectional study in three phases in different study settings (Additional file 1).

The three main phases were item development, scale development, and scale evaluation.

### Items generation

Items generation and domain identification were conducted through a literature search, community-based focus groups and healthcare expert interviews. This step was conducted with two primary purposes: (1) to define the construct clearly and (2) to determine if measures of the construct (or related constructs) already exist. The literature review identified three assessment tools based in the United States, the United Kingdom and South Korea (17–19) as the eligible reference scales due to simple language, applicability to the health system, and free availability in the open sources. The technical aspects of care were not included as the responses could be highly biased by the patient’s knowledge of illnesses. Focus group interviews were conducted to identify the patient’s point of view on principal primary care concepts and their application. The researchers reviewed all items and scaled them under four principle domains in primary care: first-contact care, continuous care, Comprehensive care, and coordinated care.

### Content and face validity

Two community physicians with MDs, one public health expert with a PhD, four family medicine experts with MDs and an academic with a Master in Philosophy participated in the content and face validation of the SL-PCAT. The initial draft (SL-PCAT-rv-0) was emailed to these seven experts to rate the domains and items based on their relevance and representativeness in the Sri Lankan healthcare system. Using the content validity ratio (CVR) (20), the experts rated each item on a 4-point scale: 4=highly relevant, 3=quite relevant or highly Relevant, but needs rewording, 2=Somewhat Relevant, 1=not relevant (21,22). If all panellists agree that an item is ‘’highly relevant”, the CVR criteria for retaining was 0.99. When the number of panellists rating an item ‘’highly relevant’’ is more than half but less than all, the CVR is between 0 and 0.99. If no raters mark the item as ‘’essential,’’ the CVR would be 0. Following the suggested revisions, e.g. rephrasing new items for clarification, the items were brought to the Delphi method session through an email thread (23). After examining the response options, the Likert scale was selected as the suitable response scale for the questionnaire.

### Items translation

Items translation from English to Sinhala, the national language spoken by 87% of the population (24), was done by a native Sinhalese speaker. The principal investigator observed the translation process to ensure clarity. An English language expert did backward translations. Any discrepancies between the original and the English version SL-PCAT-rv-1(Additional file 2) were discussed through reconciliation between translators and the principal investigator.

### Scale development

### Cognitive interviews

Cognitive interviews were conducted among representatives of the targeted population to ensure clarity in Sinhala language and technical soundness. Four individuals with varied language competencies were invited (a housewife, a clerk, a teacher, and a language expert with a PhD in Sinhala). The items in the SL-PCAT-rv-1 were guided to be rephrased in the participants’ words.

### Pilot test

Scale’s feasibility was tested by training five interviewers with prior experience in patient interviews by administering the tool to 25 randomly selected individuals (15 villagers and 10 patients attending a rural GP.

### Scale Evaluation

SL-PCAT-rv-2 (Additional file3) in the Sinhalese language included revised items on each core domain of primary care similar to other available PCATs worldwide (8–10,18). Through 28 items, it measured five core domains of primary care: first contact, comprehensive care, contextual care at individual, family and community level, coordination of care and continuous care. Each item was answered on a 4-point Likert-type scale, arranged in reverse order to prevent response bias. The Likert responses were ‘1= I can agree with that statement most of the time’, ‘2= I can agree with that statement to some extent’, ‘4 = I cannot agree with that statement sometimes’, ‘5 = I cannot agree with that statement most of the. A possibility to respond was added: ‘3 = I cannot say anything about that statement’.

### Construct validity

The construct validity step was assessed through a descriptive cross-sectional household survey exploring the primary care utilisation patterns in the district. SL-PCAT-rv-2 was distributed for self-administering among the participants. The household survey was estimated to cover 720 households in 34 district villages. The tool was administered among household participants instead of patients at health facilities to minimise response bias. The household participants were selected based on whether they had resided within the Anuradhapura district for the last three months. In the selected household, the respondent would be either the household head or an adult aged more than 18 who is taking part in healthcare-related decisions. The Sinhala language speaking and understanding were also considered.

### Analysis

A midscale value of 3 was assigned for “I cannot say anything about that statement.” The questionnaires with more than 50% missing data for the SL-PCAT-rv-2 scales were excluded in the beginning. First, a correlation analysis was performed in the responses set to evaluate whether the items had sufficient correlation. In the subsequent step, the 633 responses were split into two random subsamples, each containing 50% of the sample for performing exploratory factor analysis or EFA (Subsample one, n=320) and confirmatory factor analysis or CFA (Subsample two, n=313).

### Exploratory Factor Analysis (EFA)

Before performing the principal component analysis (PCA), the suitability of data for factor analysis was assessed with the Kaiser-Meyer-Olkin (KMO) and Bartlett’s (1954) Test of Sphericity. KMO sampling adequacy is a statistic that indicates the proportion of variance among the scale variables caused by underlying factors, and higher values (close to 1.0) generally indicate that factor analysis may be helpful with the data (25). Bartlett’s (1954) Test of Sphericity should reach the statistical significance of the chi-squared test for the null hypothesis to support the factorability of the correlation matrix (26). PCA is a data reduction technique that simplifies the complexity of high-dimensional data to smaller factors while retaining trends and patterns (27,28). The ‘Direct Oblimin’ oblique rotation was used in PCA because it assumes the presence of correlations between the factors (29,30).

Several other techniques were used to assist in deciding the number of factors to retain: Kaiser’s criterion and Scree test. In Kaiser’s criterion, only factors with an eigenvalue of 1 or more were retained for further investigation. The eigenvalue represents the total variance explained by the factor. In Catell’s (1966) scree test, all factors above the elbow or break in the plot were retained as these factors contributed most to the explanation of data (31).

The suitability of items was assessed based on the rule of 0.4-0.3-0.2, which recommends that satisfactory variables (a) load onto their primary factor above 0.40, (b) load onto alternative factors below 0.30, and (c) demonstrate a difference of 0.20 between their primary and alternative factor loadings (32). Empirical and conceptual relevance was also considered while assessing the problematic items for removal.

Cronbach’s alpha and item-total correlation assessed reliability to test internal consistency. A minimum Cronbach’s alpha value of 0.7 is accepted as adequate for a scale to be considered sufficiently reliable (33).

Likert scaling assumptions assessed the equal item convergence through the range of item-total correlation; domain score reliability through Cronbach’s alpha; item-convergent validity through item-scale correlations (minimum 0.3); and item-discriminant validity using scaling success rate (correlation of each item with other items within the same scale being more significant than with items from different scales). Construct validity is also measured through convergent and discriminant validity.

### Confirmatory Factor Analysis (CFA)

SL-PCAT-rv-3 with the finalised factor structure underwent CFA in R software (Lavan 0.6.17.) The Subsample-2 was subjected to structural equation modelling (SEM). The model’s overall goodness of fit was assessed using a combination of indices: the chi-squared test, the root mean square error of approximation (RMSEA), and an incremental fit index, the comparative fit index (CFI). After a few theoretically accepted modifications, fit indices were reassessed.

Total mean scores were analysed after allocating nominal values for the Likert responses: (5 = I can agree with that statement most of the time; 4 = I can agree with that statement to some extent; 3 = I cannot say anything about that statement; 2 = I cannot agree with that statement sometimes; 1 = I cannot agree with that statement most of the times. Descriptive statistics were performed for the data set of SL-PCAT-rv-3, and mean, SD, range, skewness, and kurtosis were assessed.

## RESULTS

### Items generation

For the initial draft of the SL-PCAT-rv-0, 25 items and four domains (accessibility of the first contact health care provider, provision of comprehensive care services, health care provision in the context of person, family and community, continuous care, and coordinated care) were identified as suitable for the development of SL-PCAT. Some of the experts’ review suggestions were to use simplified and familiar terms to ask the questions. “Family doctor” was not a well-used term for state primary care doctors. The community participants preferred identifying the family doctor as the usual or regular healthcare provider for acute healthcare needs. The terms ‘health promotion’, ‘coordination, ‘Non-Communicable Diseases’, and ‘communicable diseases’ were instructed to modify with a few examples. Broad terms such as ‘basic healthcare needs’ were suggested to be omitted due to the limited specificity of the range of services.

### Translation of the items

The experts had an extended discussion on the terms translated into Sinhala, and they suggested creating a very simplified version of the SL-PCAT-rv-1. This resulted in adding simplified items so the responders could easily understand, even without good healthcare knowledge and literacy skills.

### Cognitive interviews and pilot test

Ambiguous and misinterpreted terms were further simplified. Likert scale responses in the SL-PCAT-rv-2 were modified following cognitive interviews to be more understandable to lay persons. The initial Likert Scale of “I strongly agree with this statement” to “I strongly disagree with this statement” was modified to indicate the degree of agreement for the particular statement, Eg: “I can agree with that statement most of the time”. The pilot testing of the revised SL-PCAT-rv-1 took 20 minutes to complete by self-administration compared to the 30 minutes of interviews. The participants confirmed the scale’s readability and the scale’s self-administration.

### Scale evaluation with cross-sectional household survey in Anuradhapura district

The minimum expected number of participants per item in a scale undergoing exploratory factor analysis was 5:1 (n=140), and confirmatory factor analysis was 10:1 (33). The household survey met the minimum required sample size for factor analysis. The SL-PCAT-rv-2 was distributed among household representatives from 32 villages in the district. Out of 658 household visits, 15 (2.3%) declined participation in the study. After excluding responses with more than 50% missing values (n=10), 633 responses were eligible for the EFA.

### Socio-demographic characteristics of the study participants

The population from urban areas represented 16% (n=103) of responses. 82% (n=520) of the respondents were primarily responsible for making decisions related to health in their families. Most study participants were females (53%) and educated up to grade 11 (Table 1).

**Table 1.** The Socio-demographic characteristics of the total study population (n=633)

|  | Total (n= 633) | Subsample 1 (n=320) | Subsample 2 (n=313) |
| --- | --- | --- | --- |
|  | N % | N % | N % |
| **Age** |  |  |  |
| 18 – 64 | 523 (83) | 258 (80) | 265 (85) |
| 65< | 108 (17) | 62 (20) | 48 (15) |
| **Sex** |  |  |  |
| Male | 287 (47) | 148 (46) | 139 (44) |
| Female | 336 (53) | 169 (53) | 163 (52) |
| **Educational status** |  |  |  |
| Never schooled | 17 (3) | 10 (3) | 7 (2) |
| Up to Grade 5 | 114 (18) | 60 (19) | 54 (17) |
| Up to Grades 11 | 376 (59) | 181 (57) | 195 (62) |
| Up to grade 13 | 98 (16) | 56 (18) | 42 (13) |
| Completing/ completed<br>higher education | 17 (3) | 8 (3) | 9 (3) |
| **Civil status** |  |  |  |
| Single | 111 (16) | 53 (17) | 58 (19) |
| Married/ living together | 444 (70) | 222 (69) | 222 (71) |
| Divorced | 12 (2) | 8 (2.5) | 4 (1) |
| Widowed | 28 (4) | 17 (5) | 11 (4) |
| **Employment status** |  |  |  |
| Unemployed | 202 (32) | 105 (33) | 97 (31) |
| Employed | 260 (41) | 125 (38) | 142 (45) |
| Retired | 74 (12) | 44 (14) | 30 (10) |
| **Type of usual primary care**<br>**doctor** |  |  |  |
| State |  |  |  |
| PMCU | 43 (7) | 17 (6) | 21(8) |
| **DH** | 135 (23) | 70 (22) | 59 (21) |
| **OPD BH** | 28 (4) | 14 (4) | 12 (5) |
| **OPD TH** | 38 (6) | 20 (6) | 16 (6) |
| **OPD Ayurveda** | 1 (<1) | 1 (<1) | - |
| **Private** |  |  |  |
| **GP** | 372 (59) | 190 (59) | 156 (59) |
| **OPD Private hospital** | 15 (2) | 8 (3) | 4 (2) |
| **Ayurveda private practitioner** | 1 (<1) | - | - |

PMCU Primary Medical Care Unit DH divisional hospital OPD Outpatient department BH base hospital
TH Teaching Hospital GP general practitioner

This table presents the demographic and healthcare access characteristics of the study sample (n = 633), divided into two subsamples: Subsample 1 (n = 320) and Subsample 2 (n = 313). Each category includes frequency (N) and percentage (%) values.

Median age of a participant was 48 (IQR=38-61) years. Median monthly income was Rs. 39 000 (IQR=30 000-50 000). The median relationship duration with the doctor was 5 years (IQR=3-10). The last doctor visit occurred before two weeks (n=196, IQR=1-2) to two months (n=375, IQR=1-4), indicating 90% of the household visits to a primary care doctor in the last two months. A total of 164 (26%) of the study participants suffered from a chronic disease, and 15 participants did not know if they had any.

### Exploratory factor analysis with Principal Component Analysis (PCA)

EFA was performed in the Subsample one (n=320). The Kaiser-Meyer-Olkin value was .811, and Berlet’s Test of Sphericity reached statistical significance (X (378) =2820, p< 0.001, supporting the factorability of the correlation matrix. The communalities of the items were all above 0.3, indicating common variances among the items. The initial output of PCA revealed the presence of nine factors/components with eigenvalues exceeding 1, explaining 23.4%, 8.8%, 7.3%, 5.7%, 5.0%, 4.6%, 4.4%, 3.7% and 3.5% of the variance respectively. After the subsequent PCA, a four-factor solution was found (Figure 1).

**Fig 1.** The scree plot

The x-axis represents component/factor numbers, while the y-axis shows eigenvalues, which indicate the variance explained by each component.

The four-component solution explained a total variance of 66.8%, with components 1-4 explaining 33.8%, 13.8%, 11.9% and 7.1% of the variance, respectively. Direct oblimin rotation was performed to aid in interpreting these four components. The item-factor loadings above .45 were retained. The rotated solution revealed the presence of a simple structure (Thurstan, 1947), with four components showing several strong loadings in each component. The final four components/factors were identified as “care in the context of family and community with continuity‘, ‘accessibility’, ‘patient-centred care’ and ‘comprehensive and coordinated care’. ‘Care in the context of family and community with continuity’ included four of eleven original items from the two related domains. ‘Accessibility’ was derived from four of eight items of the conceptual domain. “Patient-centred care’ domain derived from the conceptual domain “health care provision in the context of person, family and community”. “Comprehensive and coordinated care’ formed one factor with the elimination of six of nine original items, primarily due to cross-loadings (n=4). The item reduction process and reasoning behind SL-PCAT-rv-2 are demonstrated in Additional file 4.

The component correlation matrix did not observe significant correlations between components, but a slightly negative correlation was reported between factor three (patient-centred care) and other components in Table 2. This table represents the correlation matrix for four components in statistical analysis. Each cell shows the correlation coefficient between two components, indicating the strength and direction of their relationship.

**Table 2.**
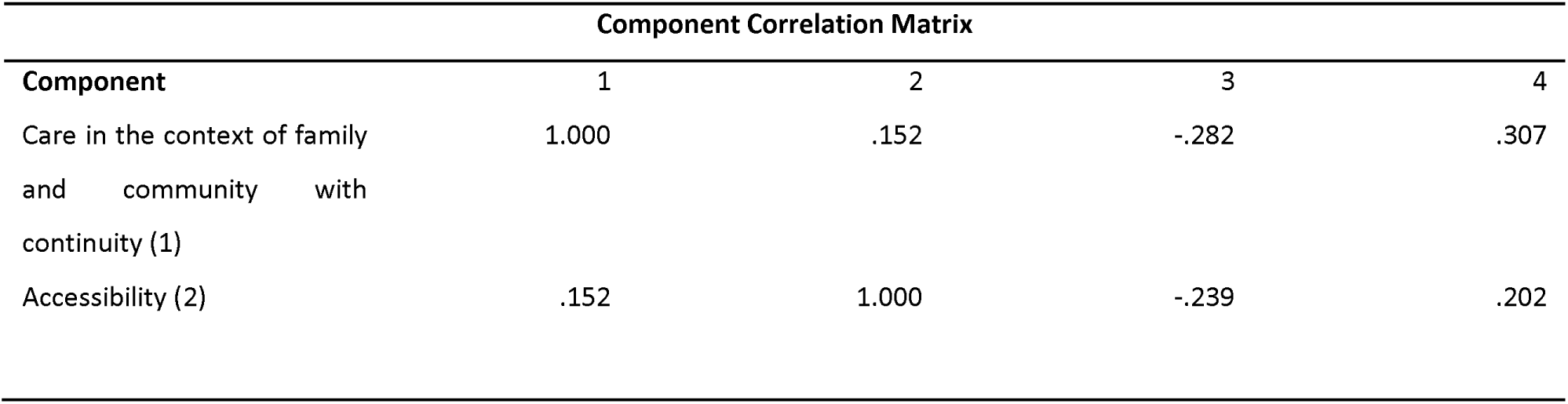
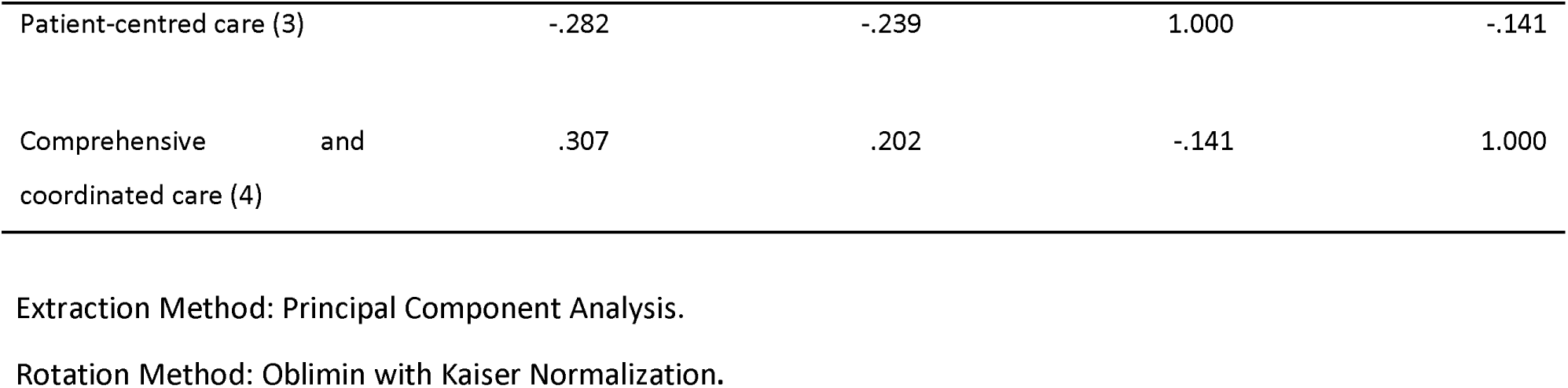
Component correlation matrix.

The final 14 items loading into the four factors in the pattern matrix are demonstrated in Additional file 5.

### Likert Scale Assumptions

All scale items showed good convergent validity because all item-scale correlations were above the minimum acceptable level, and the items in these factors were more correlated within the factor than other items, as shown in Table 3.

**Table 3.** Likert scaling assumptions using the 14 revised multi-item scales of SL-PCAT.

| Components | No of items | Item-scale correlations | Discriminant validity |  | Cronbach's alpha |
| --- | --- | --- | --- | --- | --- |
|  |  |  | Item-other scale correlation | Scaling success rate |  |
| Care in the context of family and community with continuity | 4 | 0.379-0.830 | 0.015 -0.465 | 24/37 (65%) | 0.823 |
| Accessibility | 4 | 0.331-0.461 | 0.015 -0.362 | 39/40 (98%) | 0.737 |
| Patient-centred care | 3 | 0.315-0.770 | 0.052 -0.386 | 24/30 (80%) | 0.859 |
| Comprehensive and coordinated care | 3 | 0.411-0.557 | 0.158 -0.465 | 23/33 (70%) | 0.719 |

PCD primary care doctor

This table presents the psychometric properties of four key healthcare components, assessed through various statistical measures: number of Items: Indicates the number of survey items used to assess each component; item-scale Correlations: Shows the range of correlation values between individual items and their respective components, reflecting measurement consistency; discriminant validity: displays correlation values between items from different components, ensuring they measure distinct aspects of healthcare quality; scaling success rate: represents the percentage of items that correctly align with their intended component, validating the scale’s effectiveness; Cronbach’s Alpha: measures internal consistency reliability, with values above 0.7 indicating strong reliability of the component’s measurement.

Three of the four components achieved more than 70% scale success, and most item-other scale correlations remained below 0.3, indicating good discriminative validity. The factor on ‘care in the context of family and community with continuity’ had more items correlated with factor four (comprehensive and coordinated care) and factor three (patient-centred care). However, this component was retained due to its sufficient reliability and conceptual relevance. A narrow range for item-scale correlations was observed in the ‘accessibility’ (0.13) and ‘comprehensive and coordinated care’ (0.146) components. For all revised item scales, Cronbach’s alpha coefficients ranged from 0.719 to 0.859.

### Confirmatory factor analysis (CFA)

The minimum required sample for CFA was 140 (1:10) (33). SL-PCAT-rv-3, with fourteen Items finalised through the EFA, was subjected to CFA. After allowing covariance between a few variables as suggested by modification indices, the fit measures improved to an excellent fit to data. The final chi-squared was 199, p<0.001. Absolute fit indices reported RMSEA of 0.079, GFI of 0.916 and SRMR of 0.066. Relative fit indices reported a CFI of 0.916. The finalised and revised version of SL-PCAT-rv-3 consisted of 14 items divided into four components (SL-PCAT-v-1).

### Descriptive statistics

A full range of possible scores was observed for central tendency, dispersion, and other features of the four new domains in SL-PCAT-rv-3, as demonstrated in Table 4.

**Table 4.**
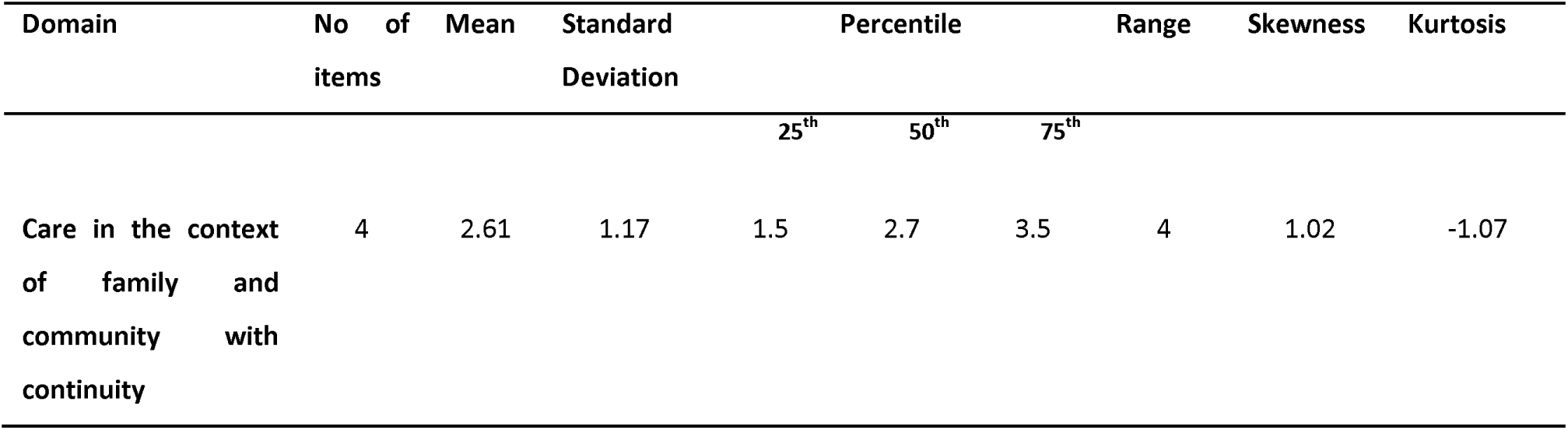
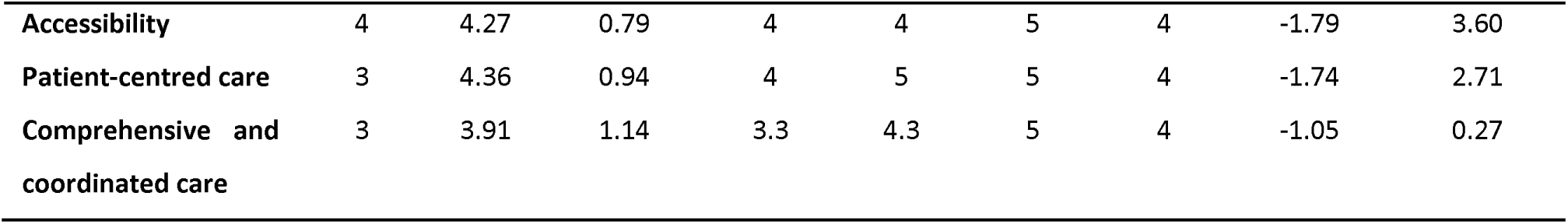
Descriptive features of SL-PCAT-v-1.

**Descriptive features of SL-PCAT-v-1**
| Domain | No of items | Mean | Standard Deviation | Percentile |  |  | Range | Skewness | Kurtosis |
| --- | --- | --- | --- | --- | --- | --- | --- | --- | --- |
|  |  |  |  | 25<sup>th</sup> | 50<sup>th</sup> | 75<sup>th</sup> |  |  |  |
| Care in the context of family and community with continuity | 4 | 2.61 | 1.17 | 1.5 | 2.7 | 3.5 | 4 | 1.02 | -1.07 |
| Accessibility | 4 | 4.27 | 0.79 | 4 | 4 | 5 | 4 | -1.79 | 3.60 |
| Patient-centred care | 3 | 4.36 | 0.94 | 4 | 5 | 5 | 4 | -1.74 | 2.71 |
| Comprehensive and coordinated care | 3 | 3.91 | 1.14 | 3.3 | 4.3 | 5 | 4 | -1.05 | 0.27 |

This table presents statistical measures of four healthcare domains, assessing their distribution and variability in a sample dataset. Key Components include the number of items; mean (the average score for each domain; Standard Deviation (SD): The variation in responses-higher SD indicates more significant variability; percentile values (25th, 50th, 75th), Showing the distribution of responses; the 50th percentile (median) represents the midpoint value; range: The difference between the lowest and highest recorded scores; skewness: measuring the asymmetry of data distribution (negative values indicate a left-skewed distribution); kurtosis: Indicating whether the distribution is more peaked or flat compared to a normal distribution. ‘Care in the context of family and community with continuity’ reported a mean of 2.61 (indicates the average score is relatively low, suggesting moderate satisfaction) and SD of 1.17 (shows variability in responses). ‘Accessibility’ reported a mean of 4.27 (high average score, indicating high satisfaction) and an SD of 0.79. ‘Patient-centered care’ reported a mean of 4.36 (high average score, indicating high satisfaction) and SD of 0.94. ‘Comprehensive and coordinated Care’ reported a mean of 3.91 (a relatively high average score, indicating reasonable satisfaction) and an SD of 1.14.

There was a significant difference in the mean scores of all four domains between state and private sector primary care doctors in the current study setting: ‘Accessibility’ = [t [315] =-5.8, p<0.001]; ‘Comprehensive and Coordinated Care’ [t(314)=-2.38, p=0.018]; ‘Patient-centered Care’=[(t [315] =-8.15, p<0.001); ‘Contextual care at the Family and Community with Continuity’= [t(314)=-2.04, p=0.04].

## Discussion

This study would be the first in Sri Lanka to develop and validate a tool for assessing patient experiences with primary care delivered by primary care doctors. SL-PCAT-v-1 comprises 14 items under four main domains: ‘Care in the context of family and community with continuity’, ‘accessibility’, ‘patient-centred care’ and ‘comprehensive and coordinated care’. The SL-PCAT-v-1 is adjusted for self-administration, increasing users’ applicability and understandability. The factors identified in the study are consistent with other validated primary care assessment tools (8,10,18). The extended conceptual and psychometric evaluations revealed this tool as feasible and reliable for measuring patient experiences.

SL-PCAT-v-1 differs from other versions of PCATs in that it has fewer items. It combined existing conceptual domains to reflect unique primary care practices in the local health system. Compared with referred versions of tools during the development process, the SL PCAT has the minimum number of items and has more similarities to the Korean version of PCAT. From the core Starfield principals of primary care (34), other than accessibility, latent domains were observed in SL-PCAT-v-1, e.g., ‘contextual care at the family and community with continuity’, ‘patient-centred care’ and ‘comprehensive and coordinated care’. Integration of family-centeredness with continuous care during the EFA was also observed in previous validation studies (10). Accessibility is assessed as a separate domain (8,9) compared to derivative domains such as ‘primary care utilisation’ and ‘economic and demographic accessibility’ (10,18). ‘Patient-centred care’ is a critical domain reflecting the care delivery to patients’ demands (10). The affordability items under ‘accessibility’ in SL-PCAT-rv-1 were eliminated during EFA due to low factor loadings and cross-loadings. Most items concerning the range of services and communication abilities of the doctors were eliminated during the EFA, and the rest were loaded into the new domain, ‘patient-centred care’.

During the assessment of the construct validity of the tool, none of the factors achieved 100% discriminative validity, indicating the lack of awareness of the public on specific services and characteristics of healthcare providers (35), which has led to misperceptions about the primary care services and challenges on assessments. The domains of SL-PCAT-v-1 reflect the varied services the local primary care doctors provide, e.g., preventive and curative care provision by state and private primary care doctors (2). Due to the high demand for care, specific primary care functions such as health promotion, immunisation, disease screening and antenatal care are currently performed by community-oriented services led by the Medical of Health (MOH).

The principal factor recognised in PCA was the ‘care in the context of family and community with continuity’. Most participants were dissatisfied with primary care doctors’ commitment to family and community-orientated care and participation in community-based programs. State healthcare authorities should adjust policies to educate the medical community about family-oriented primary care (36). Strengthening primary healthcare through comprehensive community-based and family-focused care has been a suggestion to address current healthcare service gaps (13). Public healthcare workers like Public Health Midwives deliver family-oriented care but do not fulfil the curative aspects of GPs and OPD doctors (37). Family members fulfil various healthcare needs from multiple healthcare providers, which disrupts the long-term care and the doctor-patient relationship. Most Western countries practice family-centred care extensively within their GP system (38). Family-oriented services improve health outcomes through a holistic approach, enhancing patient and family experience (39) and improving cost-effectiveness by modifying psychosocial factors through longitudinal and comprehensive care (40). This study recommends a more extensive approach to address the demand for family-oriented primary care from the local doctors through undergraduate and post-graduate training of the doctors and improving provisions for services by financial and infrastructure supports.

### Limitations

Recalling bias was the main limitation of studies that use self-administering surveys to evaluate patient experiences. Socially desirable responses concerned specific domains such as ‘contextual care at individual, family and community level’. However, to minimise the error, the study was conducted among participants from a community rather than a selected primary care facility. Most preliminary items were eliminated in the EFA process due to strict selection criteria based on factor loadings. Due to allowing modifications between variances during the CFA, over-fitting of data to the model is expected. The multifaceted nature of the health system presents a significant challenge when attempting to create a single tool that can comprehensively cover all aspects of primary care. Translations of the tool to other languages in the country require linguistic validation.

## Conclusion

The SL-PCAT scale is valid and reliable for evaluating patient perceptions of primary care delivery in Sri Lanka. The scale can gauge the implementation of fundamental primary care principles and the quality of care provided by primary care professionals. Assessing current primary care practices using the scale will help identify areas that need improvement and disparities in primary care delivery. Healthcare authorities should implement strategies to enhance primary care performance among healthcare providers, offering opportunities for improvement for primary care physicians.

## Ethical approval and consent to participate

This study was conducted in accordance with the Declaration of Helsinki, ensuring ethical principles for research involving human subjects. Ethical approval was obtained from the Faculty of Medicine and Allied Sciences Ethics Review Committee, Rajarata University of Sri Lanka (Approval No. ERC/2020/66). Informed consent was obtained from all participants prior to data collection. An information sheet in Sinhala was provided to participants, explaining the study’s purpose. No incentives were offered to participants during data collection.

## Patient and public involvement

The public and patients were involved during the tool development process by participating in focus group interviews, cognitive interviews, pilot testing, and scale evaluation through a community survey.

## Consent for publication

Not applicable

## Availability of data and materials

The datasets used and/or analysed during the current study are available from the corresponding author upon reasonable request.

## Authors’ contributions

All authors contributed to the conceptualisation, methodology, investigation, data analysis, visualisation, and validation of the study findings. P.A. contributed to funding acquisition and writing the original draft. S.B.A., S.S., and P.H.G.J.P. contributed to supervising, reviewing, and editing the manuscript.

## Data availability statement

All data relevant to the study are included in the article or uploaded as supplementary information.

## Acknowledgements

The authors would like to thank the following individuals who participated as content and language reviewers in this study: Prof. Thilini Agampodi, Prof. D. Rathish, Dr. Dinusha Perera, Dr. Nilmini Dayanananda, and Dr. Vimarshani Hewageegana. We acknowledge Prof. Janith Warnsekara and Prof. Nuwan Wikramasinghe of the Department of Community Medicine, Faculty of Medicine and Allied Sciences, Rajarata University of Sri Lanka, for the support provided during the statistical analysis of the data.

## Abbreviations

CFA: Confirmatory Factor Analysis
CFI: Comparative Fit Index
CVR: Content Validity Ratio
GFI: Goodness of Fit Index
GP: General Practitioner
KMO: Kaiser-Meyer-Olkin
M Phil: Master of Philosophy
MBBS: Bachelor of Medicine, Bachelor of Surgery
MD: Doctor of Medicine
NCDs: Non-Communicable Diseases OPD Outpatient Department
PCA: Principal Component Analysis
PMCU: Primary Medical Care Units
RMSEA: Root Mean Square Error of Approximation
rv: Revised Version
SEM: Structural Equation Modeling
SL-PCAT: Sri Lankan Primary Care Assessment Tool
SRMR: Standardized Root Mean Square Residual

---

Additional file 2.pdf

Sri Lankan Primary Care Assessment Tool (SL-PCAT) - English version/draft one (SL-PCAT-rv-1)

This is the English version of the initial draft of SL- PCAT-rv1 prior to the Sinhala translation. This content underwent content review by experts in primary care and public health.

Additional file 3.pdf

Sri Lankan Primary Care Assessment Tool (SL-PCAT) - Sinhalese translated version (SL-PCAT-rv-2)

SL-PCAT-rv-2 includes Sinhala-translated, revised items on five core domains of primary care: first contact, comprehensive care, contextual care at the individual, family, and community level, coordination of care, and continuous care. It includes 28 items and a five-point Likert scale with a neutral option.

Additional file 4.pdf

The principal component analysis and item reduction process of the Sri Lankan Primary

Care Assessment Tool (SL-PCAT) - Sinhalese translated version (SL-PCAT-rv-2)

This table includes the results of the item reduction process and the rationale behind the item retention or removal during the principal component analysis of SL-PCAT-rv2.

Additional file 5.pdf

Pattern Matrix of the 14 Items Selected for Confirmatory Factor Analysis for SL PCAT VII This table presents a pattern matrix for the 14 items chosen for confirmatory factor analysis in the Sri Lankan Primary Care Assessment Tool (SL-PCAT rv3).

