## Additional file 2 for "DEVELOPMENT AND VALIDATION OF THE SRI LANKAN PRIMARY CARE ASSESSMENT TOOL (SL-PCAT) FOR EVALUATING PATIENTS’ EXPERIENCES IN PRIMARY CARE"

Department of Family Medicine, Faculty of Medicine and Allied Sciences,
Rajarata University of Sri Lanka

### The questionnaire for the evaluation of the quality of primary care services provided to health care users in Sri Lanka

This study identifies the doctor you visit for your primary health care needs in Out-Patient Department (OPD) of Government hospital or Ayurveda hospital or General practitioner as "your first contact health care provider" or "family doctor". Please answer these questions referring to your first contact health care provider.

Dr DMPLK Abeyrathna
[Pick the date]

| No | Statement | Response in 5 point Likert scale | |
|---|---|---|---|
| | | 1 | I strongly agree with the statement |
| | | 2 | I fairly agree with the statement |
| | | 3 | I don't know/ not applicable |
| | | 4 | I disagree with the statement |
| | | 5 | I strongly disagree with the statement |
| **Accessibility of the first contact health care provider** | | | |
| **1** | My health care provider/doctor fulfills most of my personal health care needs. | | |
| **2** | My doctor is easily accessible in an urgent health care need. | | |
| **3** | My doctor is providing medical care at a place that is not far from my residency. | | |
| **4** | Waiting time before meeting the doctor is not too long. | | |
| **5** | The opening time of my primary care doctor is suitable for me. | | |
| **6** | Amount of money spent for a medical consultation with the primary care doctor is not too high. | | |
| **7** | There are instances that I have skipped going to meet my doctor due to high medical expenses. | | |
| **8** | My doctor provides enough time to talk and clarify my health related problems. | | |
| **9** | My doctor explains my health issues and treatment options adequately. | | |

| | |
|---|---|
| **Provision of comprehensive care services** | |
| **10** | My doctor treats patients regardless of age, sex and severity of illness. |
| **11** | My doctor treats mental health problems as well as physical problems of patients. |
| **12** | My doctor advises me on general health promotion activities such as importance of physical activities, healthy diet, adequate water intake and good mental health, whenever he has time. |
| **13** | My doctor counsel me on importance of prevention of communicable and non-communicable diseases |
| **Health care provision in the context of person, family and community** | |
| **14** | My doctor is knowledgeable enough of myself as an individual. |
| **15** | Doctor's concern for me as a person is adequate. |
| **16** | Doctor respects me as an individual adequately. |
| **17** | My doctor considers my opinions during medical consultations. |
| **18** | The doctor asks about health and well-being of my family members as well. |
| **19** | My doctor is also concern about the surrounding environment where I live in. |
| **20** | My doctor is concerned about the wellbeing and health related events in the community as well. |
| **21** | My doctor actively participates in health promotion activities in the community. |

| | |
|---|---|
| **Continuous care** | |
| **22** | My doctor helps in making decisions related to my health. |
| **23** | The doctor arranges follow up after a consultation, if necessary. |
| **24** | My doctor is able to provide information of my previous medical consults whenever a need arises |
| **Coordinated care** | |
| **25** | My doctor recommends other health care resources appropriately. |
