## Additional file 4 for "DEVELOPMENT AND VALIDATION OF THE SRI LANKAN PRIMARY CARE ASSESSMENT TOOL (SL-PCAT) FOR EVALUATING PATIENTS’ EXPERIENCES IN PRIMARY CARE"

Principal component analysis and item reduction process used to develop SL-PCAT (Sri Lankan Primary Care Assessment Tool)

| Preliminary primary care domain | No | Item in the preliminary tool | Missing | ‘I cannot say anything on this statement’ (%) | Factor loading (component) | Comments |
| --- | --- | --- | --- | --- | --- | --- |
| Accessibility | 1 | I can easily visit my doctor once I get a sudden fever/cold-like illness. | 5 | 5 (1.6) | 0.751 (2) | Accessibility to PCD |
|  | 2 | My doctor treats patients at a place close to my residence and easy to reach. | 4 | 3 (0.9) | 0.696 (2) | Accessibility to PCD |
|  | 3 | I can see my doctor without waiting in a long queue. | 6 | 21 (6.6) | 0.744 (2) | Accessibility to PCD |
|  | 4 | My doctor's treatment center is open at a time that is convenient for me. | 7 | 8 (2.5) | 0.658 (2) | Accessibility to PCD |
|  | 5 | The cost of treatment I get from my doctor is an amount I can afford. | 40 (deleted) | 17 (5.3) |  | No significant loading (<0.4) |
|  | 6 | There have been times when I have missed seeing my doctor because of the high cost of my treatment. | 41 (deleted) | 56 (17.5) |  | Cross-loading on two factors (4 & 9) |
|  | 7 | My doctor provides me with enough time to discuss my health problems. | 6 (deleted) | 9 (2.8) |  | Cross-loading on two factors (2 &5) |
|  | 8 | My doctor explains my medical conditions and treatments well. | 3 (deleted) | 10 (3.1) |  | Cross-loading on two factors (2 & 5) |
| Comprehensive services of your usual healthcare provider | 9 | My doctor can treat most of my illnesses. | 4 (deleted) | 6 (1.9) |  | No significant loading (<0.4) |
|  | 10 | My doctor does not show any gender bias when giving treatment | 3 (deleted) | 18 (5.6) |  | Cross-loading on two factors (5 &6) |
|  | 11 | My doctor treats patients of all ages. | 6 (deleted) | 3 (0.9) |  | Cross-loading on two factors (6 & 7) |
|  | 12 | My doctor treats patients regardless of the severity of the disease. | 3 (deleted) | 29 (9.1) |  | Cross-loading on two factors (4 & 6) |
|  | 13 | My doctor treats mental health problems as well (e.g. depression, anxiety, stress, alcohol and drug addiction). | 11 (deleted) | 167 (52.2) |  | No significant loading (<0.4) |
|  | 14 | My doctor provides health advice for maintaining a healthy body weight, eating a healthy diet, drinking enough water and maintaining good mental health. | 2 (deleted) | 39 (12.2) |  | Cross-loading on two factors (1 & 6) |
|  | 15 | My doctor provides me with adequate advice on preventing the spread of communicable diseases (e.g., Dengue, chicken pox, COVID-19) and non-communicable diseases (e.g., Heart disease, high blood pressure, diabetes). | 4 | 34 (14.4) | 0.634 (4) | Comprehensive and coordinated care of PCD |
|  | 16 | My doctor advises me on how to control my medical conditions and prevent complications when they occur. | 3 | 18 (5.6) | 0.844 (4) | Comprehensive and coordinated care of PCD |
| Health care provision in the context of person, family and  Community | 17 | My doctor recognises me well whenever I go for treatment. | - | 17 (5.3) | -0.781 (3) | Interpersonal care of PCD |
|  | 18 | My doctor takes good care of me. | 3 | 13 (4.1) | -0.913 (3) | Interpersonal care of PCD |
|  | 19 | My doctor treats me with enough respect. | 3 | 9 (2.8) | -0.901 (3) | Interpersonal care of PCD |
|  | 20 | My doctor takes details of the disease from me when providing medical treatment. | 5 (deleted) | 6 (1.9) |  | Cross-loading on two factors (2 & 6) |
|  | 21 | My doctor inquires not only about my health but also about my family members' well-being. | 6 (deleted) | 33 (10.3) |  | Cross-loading on two factors (1 & 2) |
|  | 22 | My doctor also inquires about the environment in which I live. | 16 | 45 (14.1) | 0.753 (1) | Care in the context of family and community with continuity |
|  | 23 | My doctor inquires about health-related events in the community around me. | 8 | 45 (14.1) | 0.821 (1) | Care in the context of family and community with continuity |
|  | 24 | My doctor participates in 'Shramadana' campaigns and awareness programs to create a healthy society around me. | 6 | 97 (30.3) | 0.615 (1) | Care in the context of family and community with continuity |
| Continuous care provision | 25 | My doctor provides appropriate advice and guidance for future treatment related to my medical conditions (e.g., consultations from other hospitals /clinics, performing additional tests for diseases) | 1 (deleted) | 15 (4.7) |  | No significant loading (<0.4) |
|  | 26 | My primary care doctor reviews me after giving me medical advice to discuss the progress of the conditions and explain the test results. | 6 (deleted) | 24 (7.5) |  | Cross-loading on two factors (1 & 4) |
|  | 27 | My doctor collects information about patients' illnesses in medical records. | 8 | 125 (39.1) | 0.790 (1) | Care in the context of family and community with continuity |
| Coordinated care provision | 28 | My doctor refers patients to other health-related services when necessary (e.g., hospital clinics, physiotherapists, psychiatric consultations, and other specialists). | 6 | 28 (8.8) | 0.495 (4) | Comprehensive and coordinated care of PCD |
