## Additional file 5 for "DEVELOPMENT AND VALIDATION OF THE SRI LANKAN PRIMARY CARE ASSESSMENT TOOL (SL-PCAT) FOR EVALUATING PATIENTS’ EXPERIENCES IN PRIMARY CARE"

Table 2 Pattern Matrix of the 14 items selected for confirmatory factor analysis for SL PCAT VII

|  | Component | | | |
| --- | --- | --- | --- | --- |
|  | 1 | 2 | 3 | 4 |
| 1 easily accessible | .155 | .751 | -.098 | -.098 |
| 2 closer place | -.127 | .696 | -.008 | .229 |
| 3 less awaiting time | .183 | .744 | -.043 | -.205 |
| 4 opening time | -.209 | .658 | -.021 | .340 |
| 15 prevention of CD and NCD | .243 | .145 | .011 | .634 |
| 16 control of prevailing illnesses | .050 | -.005 | -.068 | .844 |
| 17 identify person as an individual | .102 | .129 | -.781 | -.107 |
| 18 care fpr the patient | -.025 | .038 | -.913 | .020 |
| 19 respect for the patient | -.057 | -.053 | -.901 | .098 |
| 22 concerns on patients environment | .753 | .107 | -.175 | .015 |
| 23 concerns on patients sorrounding community | .821 | .046 | -.108 | .048 |
| 24 doctor engages in health promotion activities in the community | .615 | -.198 | -.198 | .170 |
| 27 keeps records of patients disease information | .790 | .084 | .238 | .078 |
| 28 coordinate other health care facilities | .375 | -.002 | -.041 | .495 |
| Extraction Method: Principal Component Analysis.   Rotation Method: Oblimin with Kaiser Normalization.^a^  a. Rotation converged in 11 iterations. | | | | |
